# Breath aerosol PCR for detection of lower respiratory tract infections: Evaluation of a non-invasive face mask collector in pneumonia patients

**DOI:** 10.64898/2026.04.18.26351117

**Authors:** Katie Tiseo, Sarah Dräger, Harshitha Santhosh Kumar, Maia Alkhazashvili, Anya Hammann, Patricia Risch, Reto Willi, Tamar Mkhatvari, Christina Fialová, Christian Adlhart, Denise Szabó, Mariam Suknidze, Izolda Pachkoria, Tobias Broger, Elena Ivanova Reipold, Khatuna Varshanidze, Michael Osthoff

**Affiliations:** Avelo AG, Schlieren, Switzerland; Department of Internal Medicine, University Hospital Basel, Basel, Switzerland; National Center for Disease Control & Public Health of Georgia, Tbilisi, Georgia; Avicenna – Batumi Medical University, Batumi University Hospital (MedCenter), Batumi, Georgia; ZHAW Zurich University of Applied Sciences, School of Life Sciences and Facility Management, Institute of Chemistry and Biotechnology, Wädenswil, Switzerland; Avicenna – Batumi Medical University, Batumi, Georgia; Hightechnology Hospital Medcenter, Batumi, Georgia; Department of Internal Medicine, Cantonal Hospital Winterthur, Winterthur, Switzerland; Department of Clinical Research, University of Basel, Basel, Switzerland

## Abstract

Etiological diagnosis of lower respiratory tract infections (LRTIs) relies on sputum or bronchoalveolar lavage (BAL), which may be difficult to obtain or invasive. Exhaled breath aerosol (XBA) sampling offers a non-invasive alternative for pathogen detection. We evaluated the performance of the AveloMask, a face mask–based device designed to capture XBAs for molecular testing. In this prospective paired-sample study, hospitalized adults with pneumonia at three hospitals in Switzerland and Georgia provided an XBA sample using the AveloMask and a lower respiratory tract (LRT) specimen (sputum or BAL). XBA samples were analyzed by multiplex PCR using the Roche LightMix® panel and LRT samples were tested using the BioFire® FilmArray® Pneumonia Panel. Concordance between XBA and LRT samples was assessed using positive percent agreement (PPA), negative percent agreement (NPA), and overall percent agreement (OPA). Ninety-three participants were enrolled and 63 participants provided paired samples. AveloMask sampling identified the dominant pathogen (lowest Ct value in the LRT sample) in 40/47 LRT-positive cases (85.1%). Across all targets, PPA was 61% (95%CI, 50–72%), NPA was 100% (95%CI, 99–100%), and OPA was 95% (95% CI, 92–96%). PPA was higher for bacteria than for viruses and lower PPA was largely driven by reduced detection of low-abundance or co-infecting pathogens. In a subset analysis, AveloMask results showed substantial overlap with standard-of-care testing and could have supported antimicrobial de-escalation. Breath aerosol sampling using the AveloMask enabled non-invasive molecular detection of LRT pathogens in pneumonia cases and may complement conventional standard-of-care testing, particularly when sputum is unavailable.

## 2. Introduction

Lower respiratory tract infections (LRTIs) causing pneumonia are a leading cause of morbidity and mortality worldwide, and account for a substantial proportion of hospital admissions and antibiotic use among adults (1–3). Accurate identification of the causative pathogen is essential to guide targeted therapy, inform infection control measures, and reduce unnecessary antibiotic use. Multiplex PCR assays have improved the sensitivity and speed of pathogen detection from respiratory specimens compared with conventional culture-based methods; however, the clinical utility of multiplex PCR depends heavily on specimen quality and type. Conventional diagnostic samples for LRTIs include sputum and bronchoalveolar lavage (BAL). High-quality sputum specimens can be difficult to obtain, and BAL is invasive and resource intensive, limiting its routine use (4, 5). While upper respiratory tract swabs are easier and faster to collect, they may not reliably reflect pathogens causing disease in the lower respiratory tract (LRT) (6, 7). These limitations can result in diagnostic uncertainty, encourage empiric broad-spectrum antibiotic prescribing and may impair timely deescalation. Ongoing debate in pneumonia guidelines - including recommendations for antibiotic use in patients with confirmed viral infections - highlights the need for improved, non-invasive diagnostic approaches that enable timely pathogen detection and more individualized antimicrobial decisions (8).

Non-invasive sampling of exhaled breath aerosols (XBAs) represents a potential complement to traditional respiratory specimens. Respiratory pathogens are known to be transmitted via aerosols, and prior studies have demonstrated that viruses and bacteria can be detected in XBAs (9–12). However, much of the prior work on XBAs has either focused on surrogate biomarkers such as volatile organic compounds (VOCs), which often have low specificity, or have used large equipment in research settings that is impractical for routine clinical workflows (13–16).

The AveloMask (Avelo AG, Switzerland) is a face mask–based breath sampling kit designed to capture XBAs during normal breathing and coughing onto an integrated filter. They are subsequently transferred into a stabilization buffer for downstream nucleic acid detection. This approach offers a scalable, non-invasive complement to traditional LRT sampling, particularly when sputum is unavailable or of poor quality. In this prospective, paired-sample study in hospitalized adults with radiographically confirmed pneumonia, we evaluated the diagnostic performance of the AveloMask by comparing pathogen detection in XBA samples with that in standard LRT specimens (sputum or BAL) using commercial multiplex PCR. We assessed concordance between AveloMask and the comparator LRT samples.

## 3. Materials and Methods

### AveloMask Kit design and procedure

The AveloMask (Avelo AG, Switzerland) is a specimen collection kit designed to collect XBA samples from the respiratory tract. The processing steps are illustrated in Fig. 1. The kit consists of a face mask and a buffer tube. The inner surface of the mask contains a filter inlay that captures the aerosolized particles from exhaled breath. The filter inlay is a 75 × 75 mm electrospun fibre sheet mounted on the inside of the mask (Supplementary Methods).

**FIGURE 1.**
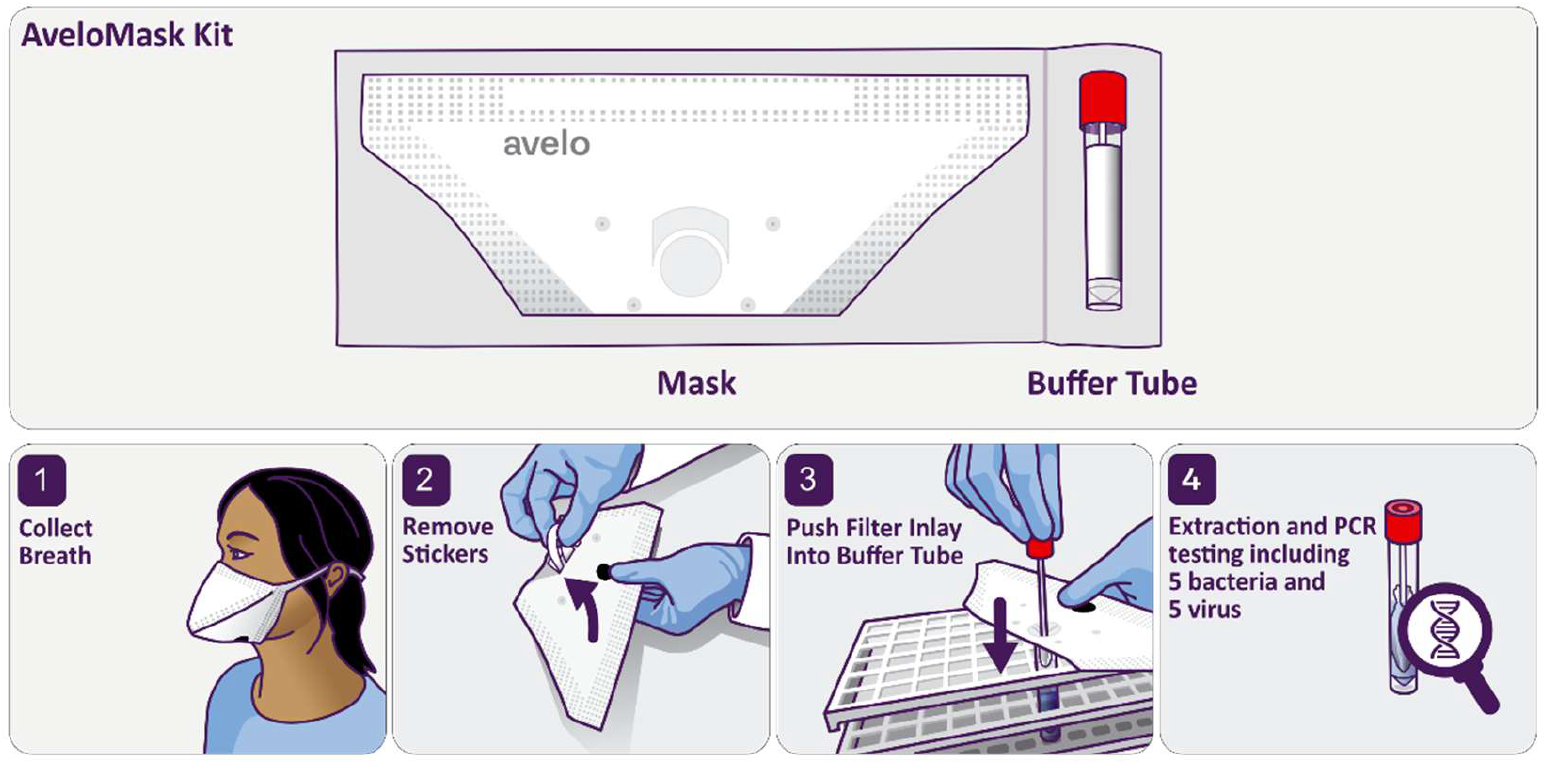
AveloMask Kit and breath aerosol collection and testing procedure. Figure adapted from Risch et al (10).

After wearing the mask (step 1), two protective stickers on either side of the mask are removed (step 2). The filter inlay is then transferred into the buffer tube by pushing it into the tube using a stick attached to the tube cap (step 3). The buffer inactivates the sample and preserves nucleic acids, allowing transport at ambient temperature until further processing or biobanking (step 4). Detailed instructions are provided in Movie S1 and Fig. S1 (10).

### Clinical evaluation

This prospective, paired-sample comparison study initially enrolled participants from an inpatient hospital site in Basel, Switzerland (clinicaltrials.gov, NCT06668883). It was subsequently expanded to include two hospital sites in Batumi, Georgia to achieve the target sample size, using closely aligned study procedures across sites. Participants were enrolled between November 13, 2024 and June 12, 2025. Eligible participants were consenting adults (≥18 years of age) with a diagnosis of LRTI defined as a new pulmonary infiltrate on chest imaging and at least one clinical symptom consistent with an LRTI, including fever, chills, cough, sputum production, dyspnoea, tachypnoea, or chest pain (Basel only). Participants were excluded if they were unable to complete all sampling procedures within 48 h of initiating new antibiotic therapy; were critically ill at the discretion of the investigator; or were receiving oxygen supplementation via face mask, high-flow oxygen, non-invasive ventilation, or mechanical ventilation. Participants receiving supplemental oxygen via nasal cannula were eligible for inclusion.

### Ethics and reporting

The protocol of this observational study was approved by the Ethics Committee of Northwest and Central Switzerland for study activities in Basel (approval no. 2024-01738) and by the Clinical Research Ethics Committee of the University Clinic of Innovative Medicine for study activities in Batumi (meeting no. PROT-UCIM-REC-001). Written informed consent was obtained from all participants prior to enrolment.

### Sampling Methods

Demographic and clinical data were collected using case report forms (CRFs). In Basel, data were recorded electronically using secuTrial, a GCP-compliant electronic data capture system (interActive Systems GmbH, Berlin (Germany)), whereas paper CRFs were used in Batumi.

Sputum collection was attempted for all participants and was performed either spontaneously or via induction according to standard clinical procedures. Sputum samples were required to appear visually mucoid or purulent at the time of collection, and samples meeting this criterion underwent further laboratory assessment. Those containing < 25 squamous epithelial cells per low-power field were considered of acceptable quality. Acceptable sputum samples were used as the comparator LRT specimen and were stored at −80 °C until shipment to Avelo AG (Switzerland) for batch testing. BAL samples were not collected as a part of this study; however, when BAL was collected as a part of routine clinical care and when no acceptable sputum sample was available, BAL samples were permitted as the comparator LRT specimen.

For breath sampling, participants wore the AveloMask for 30 min and were instructed to perform 20 deep coughs in addition to any spontaneous coughs during mask wear. Mask samples were processed on site by trained hospital staff. Filter inlays were transported in stabilizing buffer at ambient temperature on the day of collection, stored at −80 °C, and shipped on dry ice to Avelo AG for batch testing.

To gather basic feedback on ease of use, participants were asked immediately after sample collection: “Overall, how difficult or easy was it to provide the AveloMask sample?” Answers were recorded on a 5-point scale and recorded in the CRFs.

### PCR methods

#### AveloMask samples

Frozen AveloMask samples, consisting of filter inlays submerged in stabilizing buffer, were thawed and briefly centrifuged for 10 s at 2,000 rcf prior to opening to prevent aerosol contamination. The recommended amount of DNA and Equine Arteritis Virus (EAV) RNA process controls (Roche, Switzerland) and 30 µg of carrier RNA were added to each tube followed by vortexing for 10 s, and incubation in a water bath at 70 °C for 15 min. Following heat elution and lysis, the tube was vortexed for additional 10 s and centrifuged at 2,000 rcf for 10 s.

Then, the filter inlay was compressed using a disposable sterile Pasteur pipette, and the eluate was transferred to a fresh tube for nucleic acid extraction. Molecular-grade ethanol (2.0 mL) was added, and DNA and RNA were extracted from the entire sample using the QIAamp DNA Mini Kit (Qiagen, Germany) according to the manufacturer’s instructions, with omission of proteinase K and Buffer AL, as the sample buffer already contained guanidinium thiocyanate. Nucleic acids were eluted in 100 μL of Buffer AE.

Each extraction batch included a negative control consisting of buffer containing DNA and EAV RNA process controls and 30 µg carrier RNA, which was required to be negative for all pathogen targets while demonstrating appropriate amplification of process controls for run validity. For each sample, two 10-μL aliquots of extract were analysed, one for viral targets and one for bacterial targets. Quantitative PCR was performed on a LightCycler® 480 II instrument (Roche, Switzerland) using modular Roche LightMix® primer and probe sets targeting five viruses (Influenza A virus, Influenza B virus, respiratory syncytial virus (RSV), SARS-CoV-2, and rhinovirus), along with the EAV RNA process control, and five bacteria (*Mycoplasma pneumoniae, Chlamydia pneumoniae, Haemophilus influenzae, Streptococcus pneumoniae, and Legionella pneumophila*), together with the DNA process control. Separate RNA and DNA master mixes were used as required.

LightMix modular positive controls for all pathogens were included in each reaction plate. Any amplification curve consistent with a typical PCR profile, with signal sufficiently above background and a Ct value < 45 was considered positive. In cases where no pathogens were detected, process controls were checked to confirm assay validity. Samples were called invalid if both pathogens and process controls were negative. Invalid runs and samples were repeated where sufficient sample material was available. Samples remaining invalid after repeat testing were excluded from the final analysis. Laboratory personnel performing mask sample testing were blinded to participant clinical information.

#### LRT samples

Comparator LRT samples were tested using the Biofire® Filmarray® Pneumonia Plus Panel (BioMérieux, France), a CE-IVD–marked point-of-care multiplex PCR assay validated for sputum and BAL samples. This platform was selected as the reference method because it is widely implemented in routine clinical practice, is validated for LRT specimens, and provides standardized, automated results with minimal operator-dependent variability. Testing was performed according to the manufacturer’s instructions, and results were interpreted based on the automated report generated at completion of each run. In addition, 200 µL of residual LRT sample was extracted and tested using the LightMix panel following a similar protocol as for AveloMask samples. The LightMix and BioFire panels overlapped for all targets included in the LightMix panel, except for SARS-CoV-2. Laboratory personnel performing LRT sample testing were blinded to participant clinical information but were not blinded to the LRT comparator results. This reflected the centralized testing workflow and limited laboratory staffing.

#### Data analysis

Sample size was determined using a precision-based approach, targeting two-sided Wilson score confidence intervals with a width of ±15%, assuming a positive percent agreement (PPA) of 70% for mask-based qPCR relative to LRT samples across pathogens. PPA was used for sample size determination as fewer positive pathogen agreement events are expected compared to negative agreement events in the PCR panel. Assuming detectable pathogens in at least 50% of LRT samples based on previous studies (17) and an indeterminate test rate of 8% yielded a sample size of 60 participants with 55 evaluable paired XBA and LRT samples.

Point estimates and 95% Wilson score confidence intervals were calculated for PPA, negative percent agreement (NPA), and overall percent agreement (OPA) of AveloMask LightMix PCR panel results compared with LRT BioFire PCR panel results. Agreement analyses were conducted per participant, per pathogen, across all pathogens combined, and stratified by viral and bacterial targets, study site, and LRT Biofire sample Ct value. The same analyses were repeated for AveloMask LightMix PCR versus LRT LightMix PCR in a secondary analysis. We reported pathogen positivity of AveloMask versus LRT samples using heatmaps and performed an analysis to determine the percentage of participants in whom AveloMask detected the dominant LRT pathogen (defined as the pathogen with the lowest Ct value in the LRT specimen). Missing and invalid results were reported, and agreement analyses were restricted to complete paired samples with valid index and reference test results. In a subset of participants enrolled in Basel, AveloMask LightMix PCR, LRT BioFire, and available standard-of-care microbiological testing results were descriptively compared to assess pathogen detection overlap using Euler diagrams. In addition, the principal investigator performed an exploratory post hoc assessment of the potential impact of AveloMask results on antimicrobial management based on clinical information and standard-of-care test (SOCT) results available at the time of care. All analyses were performed using R (version 4.5.0) (18).

## 4. Results

### Participant characteristics

Of 122 eligible participants screened, 93 consented to participate (Fig. 2). Thirteen of these 93 participants (14.3%) were excluded because a good-quality sputum sample could not be obtained. Fourteen AveloMask samples were excluded prior to PCR analysis due to site-specific procedural deviations early in the study (n = 12) and participant-related factors (n = 2), which were subsequently addressed through additional staff training. Ultimately, 63 participants who successfully provided both an LRT sample and an AveloMask XBA sample were included. Comparator LRT specimens consisted almost exclusively of sputum (62/63, 98.4%), with one BAL sample obtained during routine clinical care. Most participants (57/63, 90.5%) had initiated in-hospital antibiotic therapy prior to sample collection. Demographic and clinical characteristics are summarized in Table 1. The median time from antibiotic initiation to AveloMask sampling was 22 h (range, −1 to 44 h). The median time interval between collection of the two sample types was 1 h (range, 2 min – 25 h). The study population primarily consisted of middle-aged adults, with a slight predominance of male participants. No adverse events or complications related to either AveloMask sampling or collection of LRT comparator specimens were reported.

**FIGURE 2.**
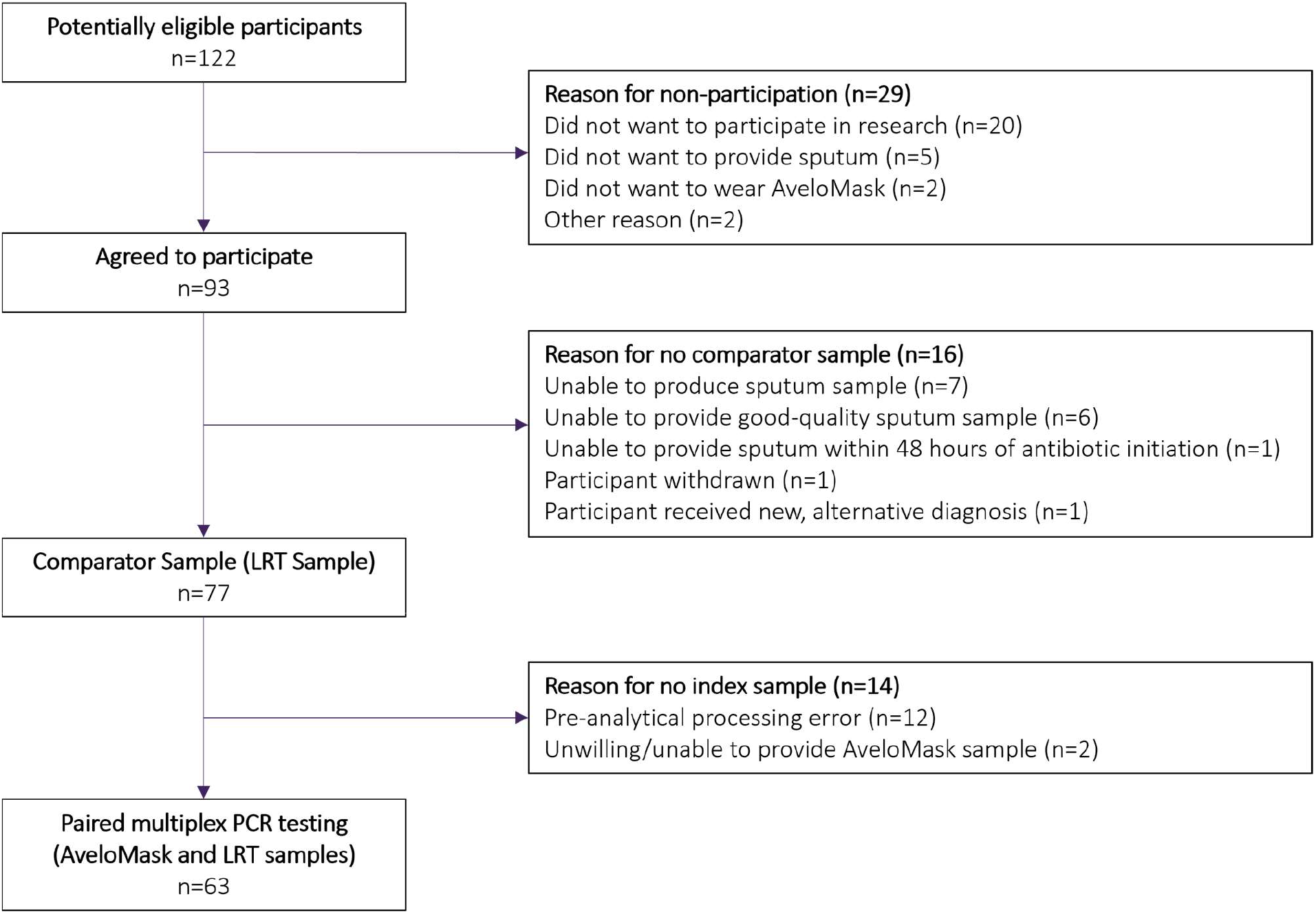
Study flowchart. Abbreviations: LRT, lower respiratory tract

**TABLE 1.** Characteristics of the study cohort. Continuous data are presented as median (range) and categorical data as counts (relative percentages). ^1^Immunocompromising condition or treatment was defined as a haematological disease, an oncological disease, human immunodeficiency virus (HIV), haemodialysis, solid organ transplantation, stem cell transplantation, currently undergoing chemotherapy, on chronic oral steroids, and/or on immunosuppressant drugs ^2^Range begins at −1 as one patient started antibiotic treatment post-AveloMask collection Abbreviations, CRP, C-reactive protein; CURB-65, confusion, urea, respiratory rate, blood pressure, age ≥ 65 years

| Characteristics | n = 63 |
| --- | --- |
| Age (years) | 63 (19 – 88) |
| Sex, Female | 23 (37) |
| Immunocompromising condition or treatment<sup>1</sup> | 11 (17) |
| **Clinical and Laboratory Findings on Admission** |  |
| Fever | 34 (54) |
| Chills | 5 (8) |
| Cough | 54 (86) |
| New or worsening sputum production | 25 (40) |
| Dyspnoea | 14 (22) |
| Tachypnoea | 9 (14) |
| CRP (mg/L) (n = 62/63) | 68 (1 – 406) |
| White blood cell count (g/L) | 10.5 (2.3 – 31.1) |
| Respiratory rate (n = 60/63) | 30 (14 – 40) |
| Oxygen saturation (% SpO<sub>2</sub>) | 90 (80 – 99) |
| Heart rate | 124 (60 – 151) |
| **Sample Timing** |  |
| Hours between antibiotics (if any) and AveloMask sampling (n = 59/63) | 22 (-1 – 44)<sup>2</sup> |
| **Severity Index Scores** |  |
| CURB-65 Score | 2 (0 – 4) |
| Charlson Comorbidity Index | 3 (0 – 9) |

### Per participant concordance

Overall, 47 participants (74.6%) had at least one overlapping-panel pathogen identified (Fig. 3). Considering only pathogens included on both the LightMix and BioFire panels, 23 participants (36.5%) had a single pathogen detected in their LRT sample, 24 (38.1%) had coinfections, and 16 (25.4%) had no pathogen detected. Among participants with no overlapping pathogen detected in LRT samples (n = 16), alternative microbiological diagnoses were established for 10 participants using the LRT BioFire result for non-overlapping pathogens. Identified pathogens included *Staphylococcus aureus* (n = 2), *Streptococcus agalactiae* (n = 2), Human metapneumovirus (n = 2), Adenovirus (n = 2), *Klebsiella pneumoniae* (n = 1), Rhinovirus (in a case with an indeterminate AveloMask result (n = 1)), and Human coronavirus (n = 1). In six participants no known microbiological aetiology was identified. Although SARS-CoV-2 was included in the LightMix panel, there were no positive hits in either sample type.

**FIGURE 3.**
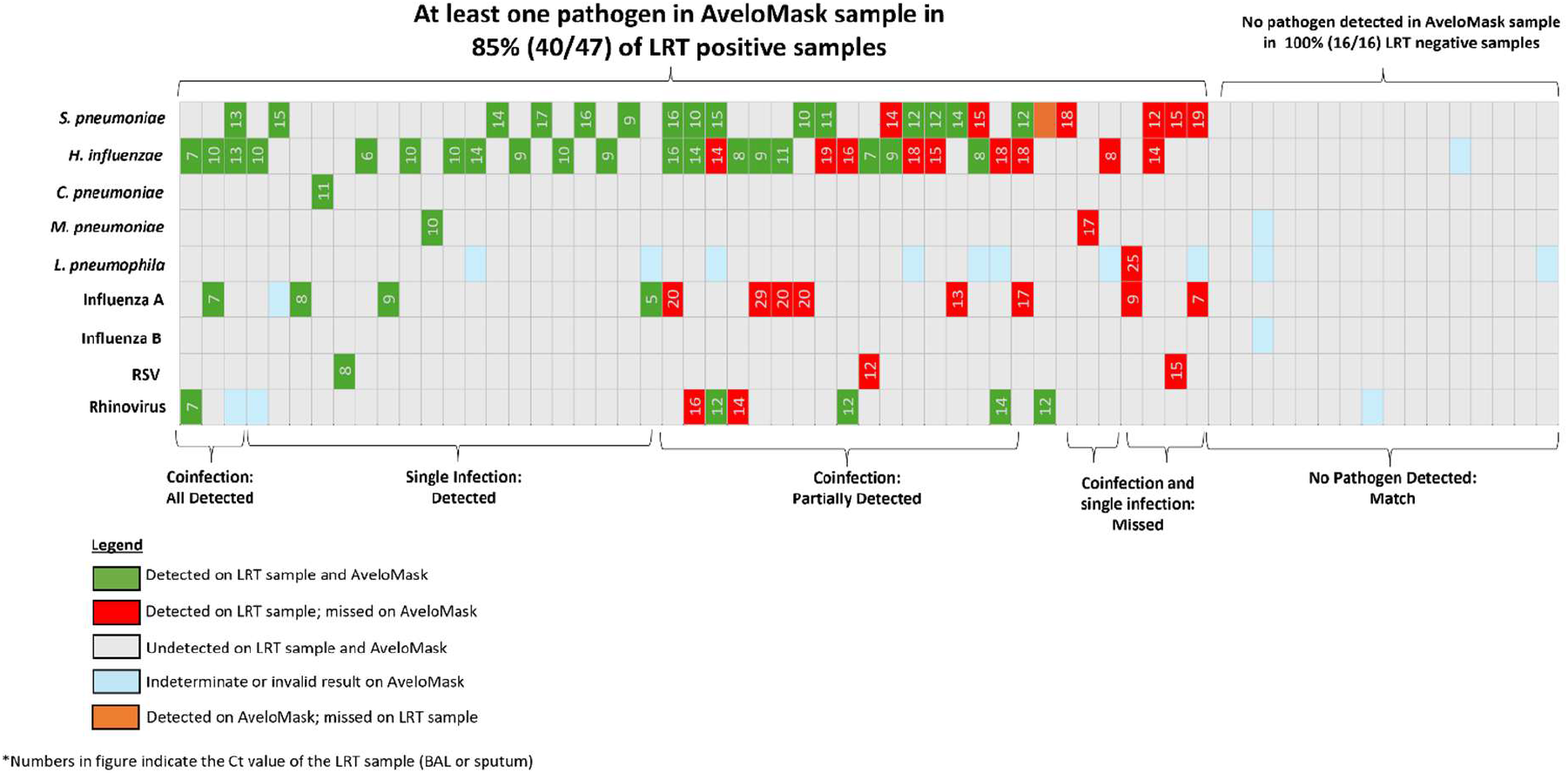
Hit map for per participant concordance between paired LRT BioFire and AveloMask LightMix PCR results. The numbers indicated in the figure show the Ct values of the LRT sample using the BioFire panel. Abbreviations: LRT, lower respiratory tract; RSV, Respiratory Syncytial Virus.

In participants with multiple pathogens detected in their LRT sample, the pathogen with the lowest Ct value in the LRT sample (and therefore highest abundance, subsequently called the “dominant pathogen”) was consistently the pathogen detected in the paired AveloMask sample. AveloMask sampling detected the dominant LRT pathogen in 40/47 (85.1%, 95% CI, 72–93%) LRT-positive participants. Among the seven participants with pathogen-positive LRT samples and no corresponding detection by AveloMask sampling, a total of 11 pathogens were identified in the LRT samples. Eight of these 11 pathogens were detected at high Ct values (> 12) in the BioFire panel, suggesting relatively low pathogen abundance. It should be noted that Ct values reported by the BioFire panel are not comparable and lower than conventional qPCR Ct values, as the assay employs a nested multiplex PCR design. One additional missed pathogen had a low Ct value (8) but occurred in the context of a coinfection with human coronavirus, which was detected at a lower Ct value and was not part of the LightMix panel used for AveloMask testing. The remaining two missed pathogens were both Influenza A virus, both detected in LRT samples at relatively low Ct values (7 and 9), with no evidence of other dominant pathogens in the corresponding LRT samples. The timing between antibiotic initiation and AveloMask sampling in these seven participants (median, 22 h; range, 0–35 h) was similar to that observed in the overall study population.

One participant tested positive for *S. pneumoniae* by AveloMask sampling but was negative for *S. pneumoniae* in their paired LRT sample. Review of available SOCT data revealed a positive pneumococcal urine antigen test and a positive sputum culture in this participant, supporting the presence of *S. pneumoniae* infection.

Overall, pathogens detected by AveloMask sampling were associated with lower LRT Ct values on the BioFire panel (mean Ct, 11), whereas pathogens not detected by AveloMask sampling had higher mean Ct values in their corresponding LRT samples (mean Ct, 16).

### Per-pathogen and overall concordance

Overall concordance between AveloMask and LRT samples are summarized in Table 2. *H. influenzae* and *S. pneumoniae* were the most frequently detected bacterial pathogens in both sample types and demonstrated PPAs of 68% and 71%, respectively. Rhinovirus was the most common viral pathogen and showed a PPA of 71%. In contrast, lower PPAs were observed for Influenza A and RSV, both at 33%. The number of RSV-positive LRT samples was limited (n = 3), whereas Influenza A was more frequently detected in LRT samples but less consistently identified in paired AveloMask samples. Across all shared targets, AveloMask sampling demonstrated an overall PPA of 61% (95% CI, 50– 72%), NPA of 100% (95% CI, 99–100%), and OPA of 95% (95% CI, 92–96%) compared with matched LRT samples. Across 563 pathogen-level comparisons (9 targets in 63 participants), 17 (3.0%) AveloMask LightMix results were invalid or indeterminate.

**TABLE 2.** Per-pathogen and overall concordance between detections for paired LRT Biofire samples and AveloMask LightMix samples (n = 63) Abbreviations: LRT, lower respiratory tract; OPA, overall percent agreement; PPA, positive percent agreement; NPA, negative percent agreement; PPV, positive predictive value; NPV, negative predictive value

| Pathogen | AveloMask<br>and LRT<br>positive | Only LRT<br>positive | Only<br>AveloMask<br>positive | Total negative | OPA | PPA | NPA | PPV | NPV |
| --- | --- | --- | --- | --- | --- | --- | --- | --- | --- |
| **Bacteria** |  |  |  |  |  |  |  |  |  |
| *Streptococcus pneumoniae* | 15 | 6 | 1 | 41 | 89% | 71% | 98% | 94% | 87% |
| *Haemophilus influenzae* | 19 | 9 | 0 | 34 | 85% | 68% | 100% | 100% | 79% |
| *Chlamydia pneumoniae* | 1 | 0 | 0 | 62 | 100% | 100% | 100% | 100% | 100% |
| *Mycoplasma pneumoniae* | 1 | 1 | 0 | 60 | 98% | 50% | 100% | 100% | 98% |
| *Legionella pneumophila* | 0 | 1 | 0 | 52 | 98% | 0% | 100% | NA | 98% |
| **Virus** |  |  |  |  |  |  |  |  |  |
| Influenza A | 4 | 8 | 0 | 50 | 87% | 33% | 100% | 100% | 86% |
| Influenza B | 0 | 0 | 0 | 62 | 100% | NA | 100% | NA | 100% |
| Respiratory syncytial virus (RSV) | 1 | 2 | 0 | 60 | 97% | 33% | 100% | 100% | 97% |
| Rhinovirus/Enterovirus | 5 | 2 | 0 | 53 | 97% | 71% | 100% | 0% | 98% |
| **Overall** | **46** | **29** | **1** | **474** | **95%** | **61%** | **100%** | **98%** | **94%** |
| Lower 95% Confidence Interval |  |  |  |  | 92% | 50% | 99% |  |  |
| Upper 95% Confidence Interval |  |  |  |  | 96% | 72% | 100% |  |  |

Subgroup analysis per site and pathogen group showed a trend towards similar PPAs in Switzerland and Georgia, and a tendency toward higher PPA in bacteria compared to virus (Tables S1 and S2). When stratifying based on low vs high Ct value with a threshold of > 12 in the corresponding LRT sample, overall AveloMask PPA reached 91% in the low Ct group and 38% in the high Ct group (Table S3). In a secondary analysis, agreement metrics were recalculated using Roche LightMix results from LRT samples as the reference standard, thereby harmonizing assay methodology between index and comparator tests. Using this approach, overall PPA was 63% (95% CI, 51–73%) and NPA was 100% (95% CI, 99–100%), with performance estimates similar to those observed when using the BioFire reference standard (Table S4).

### Comparison with standard-of-care testing

Standard-of-care testing (SOCT) data were available for 23 participants enrolled in Basel, in whom a total of 75 routine diagnostic tests were performed (mean, 3.2 tests per participant). Across AveloMask LightMix PCR, LRT BioFire, and SOCT, 34 pathogens were detected in total. Euler diagrams illustrating pathogen detection overlap are shown in Fig. S2. AveloMask detected 20/34 pathogens (59%), SOCT 18/34 (53%), and LRT BioFire 32/34 (94%). Relative to SOCT, AveloMask did not detect 8 pathogens, including 7 co-infections (6 viral [4 Influenza A, 1 RSV, 1 Rhinovirus] and 1 *S. pneumoniae*) that occurred alongside a more abundant bacterial infection. LRT BioFire detected 6 additional pathogens that were not detected by either AveloMask LightMix PCR or SOCT. AveloMask yield was higher for bacterial than viral pathogens (73% vs. 33%) (Fig. S3).

### Potential impact on treatment

In an exploratory post hoc review by the principal investigator, consideration of AveloMask results (available within 24 hours) together with SOCT data was judged to have the potential to modify immediate antimicrobial management in 30% (7/23) of patients, including 4 de-escalations (switch to penicillin/amoxicillin following detection of *S. pneumoniae*), 2 treatment discontinuations (due to lack of bacterial detection and low levels of procalcitonin), and 1 optimization (switch from beta-lactam therapy to targeted atypical coverage following detection of *M. pneumoniae*), while no immediate change was considered indicated in the remaining 16 patients. In several patients, bacterial targets detected in LRT BioFire samples were not aligned with the antibiotics prescribed, and their clinical relevance remains uncertain.

### User Feedback

A total of 83 participants attempted AveloMask sampling during the study, including individuals whose mask samples were collected prior to insufficient LRT sampling or were later excluded from PCR analysis due to processing errors. Usability was therefore assessed in all participants who underwent mask sampling. Most participants reported a positive experience: 64 of 83 (77%) rated the procedure as very or somewhat easy, 11 (13%) as neutral (“okay”), and 8 (10%) as somewhat or very difficult. Overall, AveloMask sampling was well tolerated in this population (Fig. S4).

## 5. Discussion

In this study of 63 individuals with LRTIs, XBA sampling with the AveloMask Kit followed by qPCR detection with a commercial multiplex PCR panel consistently identified the organism associated with the lowest Ct value in the matched LRT sample, indicating preferential detection of the dominant pathogen in 85% of participants. Target product profiles for LRTI diagnostics that use minimally invasive samples, such as breath, describe a clinically driven performance target of ≥ 85% sensitivity for actionable pathogen detection. The dominant-pathogen detection rate observed here meets this expert-informed benchmark for non-invasive respiratory diagnostics (19). Notably, AveloMask predominantly missed co-infecting pathogens with lower abundance in LRT samples, typically occurring alongside a more abundant bacterial infection, supporting the biological premise that exhaled pathogen abundance may reflect respiratory shedding intensity and contagiousness (20, 21). From a clinical perspective, this observation is important because identifying the predominant etiologic agent may be sufficient to guide targeted antimicrobial therapy in many cases of pneumonia (22).

More broadly, differences between AveloMask and LRT sample results may also reflect the distinction between colonization and infection, particularly for bacterial pathogens detected using the BioFire Pneumonia Panel (23). Organisms such as *H. influenzae* are frequently detected in LRT samples but may represent colonization rather than the causative agent of disease (24). In this context, the preferential detection of pathogens with higher abundance (i.e., lower Ct values) by AveloMask sampling may better reflect the dominant infecting organism, as a higher burden is more likely associated with active disease. This concept is consistent with prior work highlighting challenges in distinguishing colonization from infection in molecular diagnostics (25, 26).

Across all targets, the AveloMask Kit achieved an overall PPA of 61%. While lower than the concordance observed for dominant-pathogen detection, this reflects the preferential detection of high-abundance pathogens by AveloMask and the reduced detection of low-abundance, mostly co-infecting organisms, as well as the potential detection of less clinically-relevant pathogens by the LRT BioFire reference. This pattern suggests that discordance is more likely driven by biological factors rather than by technical failure of the AveloMask or the assay. Concordance was higher for bacterial than viral pathogens and discordance was particularly pronounced for Influenza A. This may reflect the clinical course of infection, in which viral shedding in breath has already declined by the time of breath collection at hospital presentation, while secondary bacterial infection becomes dominant. This is in line with prior studies showing that exhaled viral abundance can decrease rapidly over time (27, 28).

Recent evidence also suggests that upper respiratory tract qPCR for influenza, SARS-CoV-2, and RSV may be overly sensitive for assessing contagiousness and argues for prioritizing those with low Ct values for isolation (29, 30). When LRT results were stratified for low Ct values, PPA of AveloMask qPCR increased to 91% suggesting that AveloMask may be a better proxy for contagiousness with the potential to guide isolation practices (Table S3). If confirmed in dedicated studies, XBA sampling may help reduce unnecessary isolation and its associated economic and psychological burden.

NPA of AveloMask LightMix PCR was 100% against LRT BioFire results, with one instance of an AveloMask-positive/LRT-negative result for *S. pneumoniae*. Although the matched LRT PCR result was negative, SOCT for this participant, including a positive pneumococcal urine antigen test and clinical microbiology findings, supported the presence of pneumococcal infection. This case illustrates that breath sampling has the potential to detect clinically relevant pathogens not captured in comparator testing and underscores the imperfect nature of any single reference standard in respiratory diagnostics. While these findings suggest high specificity of AveloMask sampling, interpretation is limited by the absence of a universally accepted gold standard for etiological LRTI diagnostics and the potential for molecular testing of LRT samples to detect organisms of uncertain clinical relevance (31).

Beyond NPA, comparison with SOCT provides additional clinical context. In a subset of participants with available SOCT data, exploratory post hoc comparison of AveloMask and LRT results with SOCT showed that pathogen detection by AveloMask substantially overlapped with routine diagnostics and preferentially identified clinically relevant pathogens. In contrast, additional detections by LRT BioFire sometimes represented coinfections or organisms of uncertain clinical significance. The assessment further suggested that AveloMask results available within 24 hours could have influenced antimicrobial management in approximately one-third of cases, predominantly through de-escalation or discontinuation of antibiotics. These findings, although preliminary, suggest that breath-based pathogen detection may support more targeted antimicrobial use and complement existing diagnostic workflows. This is especially relevant in light of ongoing debates in pneumonia guidelines regarding antibiotic use in patients with confirmed viral infections, underscoring the need for improved tools to guide treatment decisions (8).

Breath-based sampling offers several potential advantages over conventional approaches. Sputum collection can be challenging in hospitalized patients, as we confirmed in this study. Of the 93 participants enrolled in this study, 13 (14%) were excluded due to issues with sputum production. These numbers are in line with other studies that show difficulty in obtaining adequate sputum samples in this population (32–34). In contrast, mask sampling was feasible in routine ward settings and was well-tolerated by older hospitalized pneumonia patients, with most participants rating the procedure as easy or very easy. Together, these findings support the potential of XBA sampling as a complementary diagnostic approach to increase diagnostic yield, particularly when sputum is unavailable or of poor quality.

Several studies have evaluated other non-invasive diagnostic approaches for pneumonia. The high NPA observed in this study is consistent with the principle of aerosol-based sampling and highly specific PCR, which detects pathogen-derived nucleic acids from the LRT. In contrast, upper respiratory tract PCR testing, while reaching comparable PPAs, may be more prone to detecting colonizing organisms and therefore tends to show lower NPAs in relation to LRT infection (35, 36). Alternative non-invasive approaches have mostly focused on host biomarkers, including VOCs and blood-based signatures, which do not provide direct pathogen identification and have shown even more limited specificity, with VOC-based approaches reaching approximately 83% specificity in a recent meta-analysis (16).

This study has several strengths. First, the paired-sample design enabled direct within-participant comparison between XBA and LRT specimens, reducing confounding from inter-individual variability. Second, participants were prospectively enrolled using standardized clinical criteria, and comparator samples were restricted to good-quality LRT specimens, ensuring a clinically well-defined cohort. Third, exclusions and invalids were systematically reported according to the Standards for Reporting Diagnostic Accuracy Studies (STARD, Table S5). Fourth, the study evaluated both performance and collected user feedback, providing insight into real-world feasibility of breath-based sampling in hospitalized patients with LRTIs. Fifth, two PCR panels, including a well-accepted and CE-marked sample-to-answer system were used for LRT sample reference testing and delivered comparable results.

This study also has several limitations. First, the study population may be subject to selection bias because inclusion required availability of a good-quality LRT comparator, which was predominantly sputum. This may limit generalizability to patients unable to expectorate, in whom the clinical utility of XBA sampling may be highest. Second, although enrolment was restricted to participants who had initiated antibiotic therapy within 48 hours, this interval may have permitted substantial reductions in XBA bacterial abundance, potentially lowering concordance between breath and LRT samples, particularly as sputum sampling was generally performed prior to XBA collection (37–39). Third, the limited number of detections for several individual pathogens reduced the statistical precision of pathogen-specific performance estimates, as reflected by wide confidence intervals. Fourth, a comparison to SOCT data was only possible in participants from one site, and assessment of performance against culture or a composite reference standard requires further studies. Finally, SARS-CoV-2 was not included in the LRT BioFire panel and was therefore assessed only in the secondary LightMix-based analysis, in which no samples were positive.

## 6. Conclusion

AveloMask breath sampling demonstrated biological feasibility for molecular detection of LRT pathogens and identified the dominant pathogen in matched LRT samples with high concordance. Overall PPA was moderate, largely reflecting reduced detection of low-abundance and co-infecting pathogens, while NPA was 100%. The non-invasive procedure was well-tolerated in hospitalized pneumonia patients, supporting breath-based sampling as a non-invasive adjunct to conventional LRT diagnostics. Larger studies and careful interpretation of XBA results in the clinical context are warranted to further define its clinical role.

## Ethics approval

Integrated in methods

## Data availability

All data generated or analysed during this study are included in the article, in the supplemental material, and supplemental data spreadsheet.

## Acknowledgements

This research was supported by Avelo Inc. (Switzerland), Innosuisse (award: 107.660 SIP-LS) and the Eurostars program (award BREATHCOUNTS 3718), co-funded by Innosuisse and the European Union. Avelo staff was involved in the design, data collection, analysis, and decision to publish. ChatGPT (OpenAI) was used for language editing and text shortening. The authors critically reviewed, edited, and verified all content and take full responsibility for the final manuscript.

## Funding

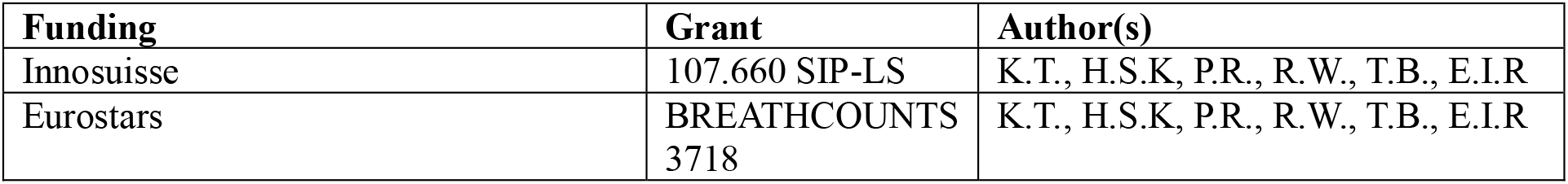

## Author contributions

S.D., K.T. A.H., T.B., E.I.R, M.A., and M.O. designed the study. D.S., A.H., T.M., M.S., I.P., K.V., M.A., S.D., and M.O. coordinated participant enrolment and clinical data collection. P.R, C.F, and H.S.K. produced mask kits. H.S.K and R.W. analysed clinical samples. K.T. performed statistical analysis. K.T. and T.B. drafted the manuscript; M.A., C.A., T.B., K.V., and M.O. provided oversight and supervision. All authors contributed, revised, and approved the final version. Authorship order follows contribution and JCM criteria.

