## Supplemental Tables and Figures for "Breath aerosol PCR for detection of lower respiratory tract infections: Evaluation of a non-invasive face mask collector in pneumonia patients"

**Table of Contents**

AveloMask Kit Production	2

Standard-of-Care Testing	2

Movie S1: AveloMask Kit Instructions	3

Figure S1: AveloMask Kit User Instructions	4

Figure S2: Pathogen Detection Overlap in participants from Basel (n=23)	5

Figure S3: Bacteria vs. Virus Detection Overlap in 23 participants from Basel	6

Figure S4: Participant Feedback	7

Table S1: Per-pathogen and overall concordance between detections for paired LRT samples (Biofire panel) and AveloMask samples (LightMix panel) stratified by study site.	8

Table S2: Per-pathogen and overall concordance between detections for paired LRT samples (Biofire panel) and AveloMask samples (LightMix panel) stratified by bacteria and virus.	9

Table S3: Per-pathogen and overall PPA for paired LRT samples (Biofire panel) and AveloMask samples (LightMix panel) stratified by LRT sample Ct value	10

Table S4: Per-pathogen and overall concordance between detections for paired LRT samples (Lightmix panel) and AveloMask samples (LightMix panel).	11

Table S5: STARD Checklist	12

**Supplementary Methods**

### AveloMask Kit Production

AveloMask Kit Production was done according to Risch et al. JCM. 2025 (<https://doi.org/10.1128/jcm.00546-25>) as follows: Filter inlays were produced via electrospinning using a Fluidnatek LE-100 system (Bioinicia Fluidnatek SLU, Spain). A 12% (w/v) solution of Polyamide 6 (Mw = 55,600 Da, BASF, Switzerland) in acetic acid/formic acid (2:1 w/w, from Carl Roth, Switzerland, and ABCR, Switzerland, respectively) was prepared and electrospun onto a spunbond polypropylene substrate following a proprietary protocol. The resulting fibre mats were cut into 75 mm × 75 mm sheets and heat-sealed along the edges using a custom-made stamping and sealing machine to form the filter inlay. The filter inlay is subsequently mounted in a releasable manner on the inner side of a custom-made duckbill face mask. Prior to inserting the filter inlay, two custom-made peel-off stickers were applied on each side of the face mask to allow processing of the filter inlay after breath collection as described in the user instructions below. Buffer tubes were produced by attaching a polyamide stick to the tube lid of standard lab tubes (Copan, Italy) and filling tubes with 3 mL of guanidinium thiocyanate buffer.

### Standard-of-Care Testing

Standard-of-care testing (SOCT) data were available for participants (n=23) enrolled at the Basel study site, whereas comprehensive SoC testing data were not routinely available for participants enrolled in Batumi. SOCT included 75 tests as clinically indicated (mean, 3.2 tests/participant) including NP/OP Swab PCR (n=22), sputum culture (n=14), BAL culture (n=3), Urine antigen test (n=18), and blood culture (n=15)

**Supplemental Figures, Videos, and Tables**

### Movie S1: AveloMask Kit Instructions

The movie shows the steps to collect and process an AveloMask sample.

### Figure S1: AveloMask Kit User Instructions

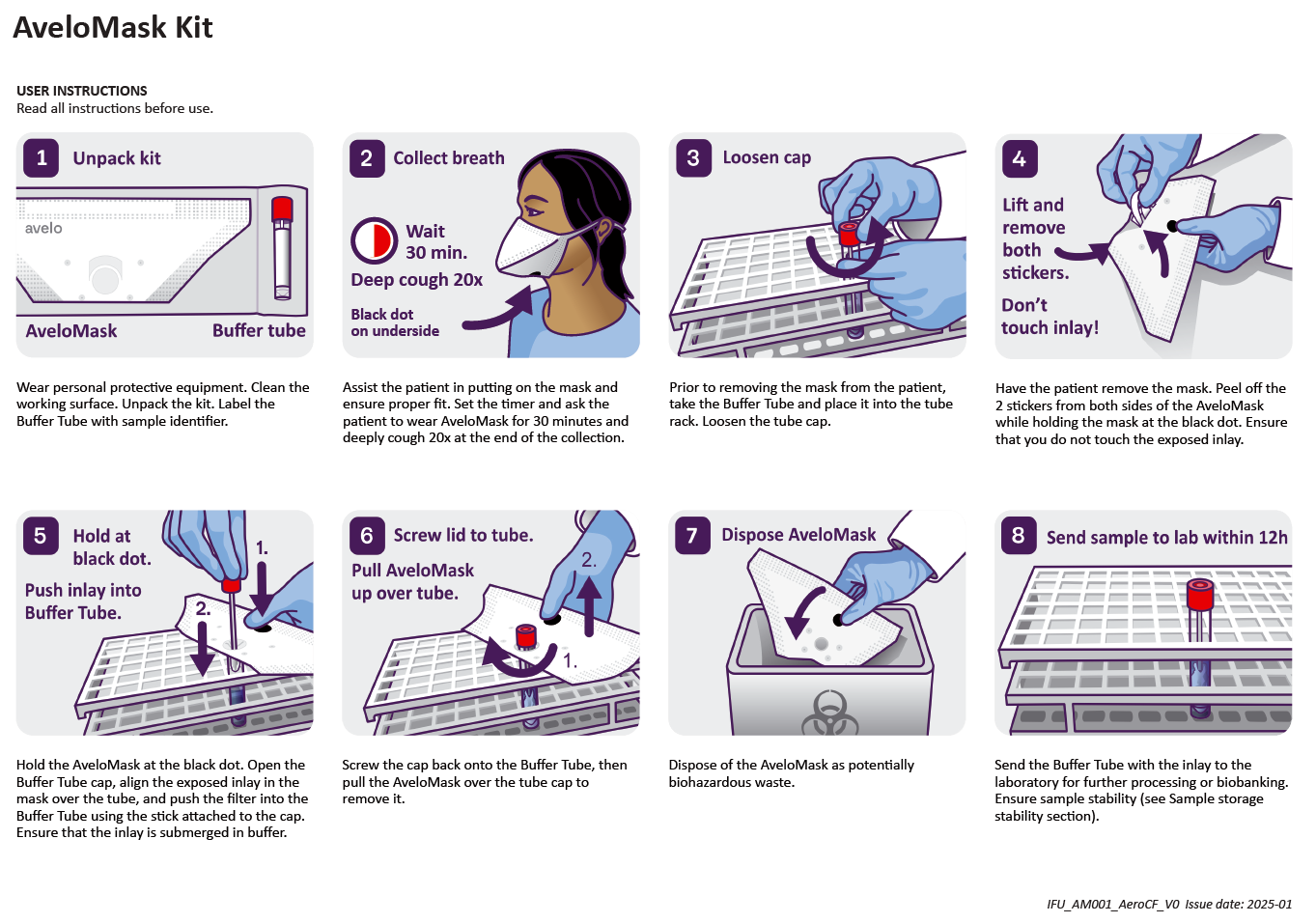

### Figure S2: Pathogen Detection Overlap in participants from Basel (n=23)

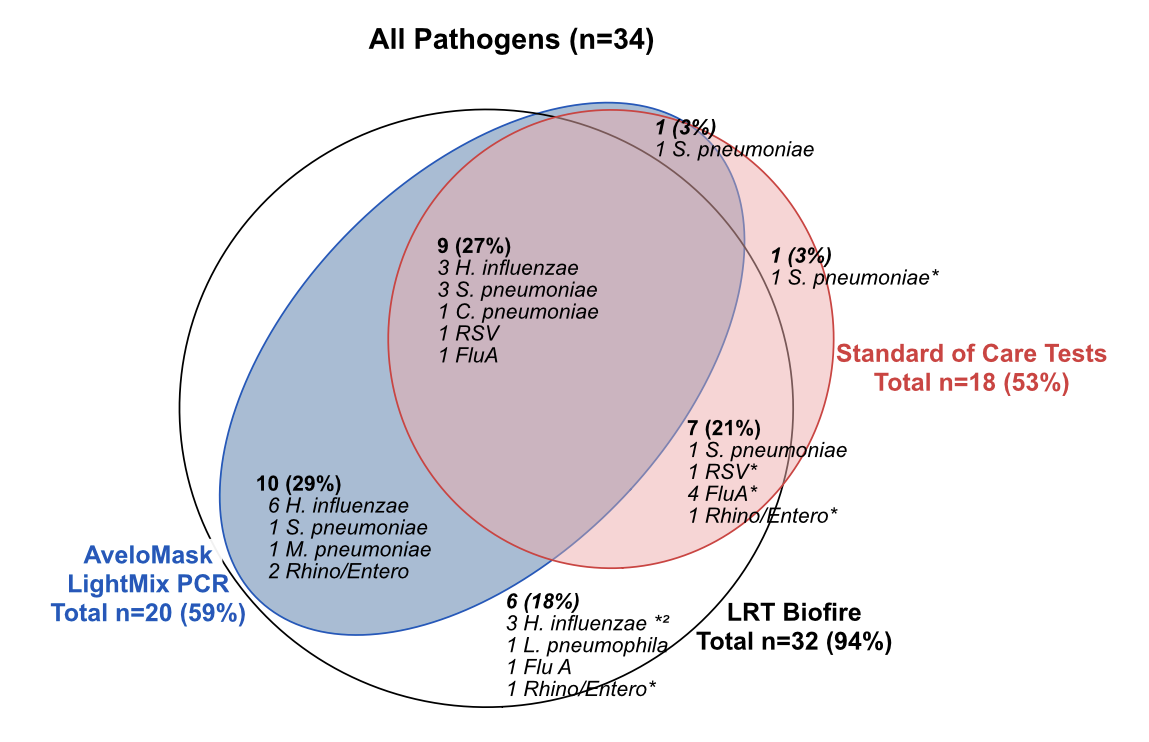

*Co-infecting pathogen not detected by AveloMask LightMix PCR in the presence of a more abundant pathogen. Standard-of-care testing included 75 tests (mean, 3.2 tests/participant) including NP/OP Swab PCR (n=22), sputum culture (n=14), BAL culture (n=3), Urine antigen test (n=18), blood culture (n=15)

### Figure S3: Bacteria vs. Virus Detection Overlap in 23 participants from Basel

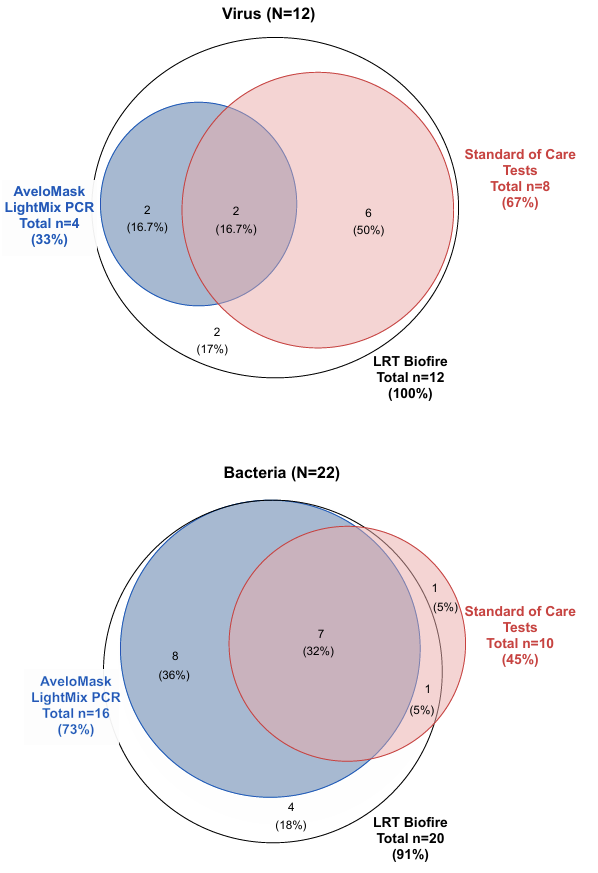

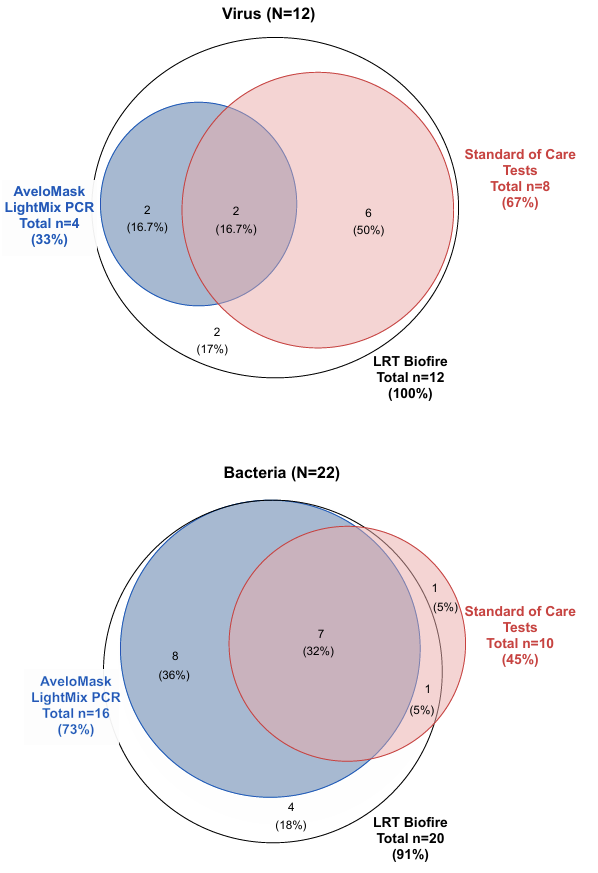

### Figure S4: Participant Feedback

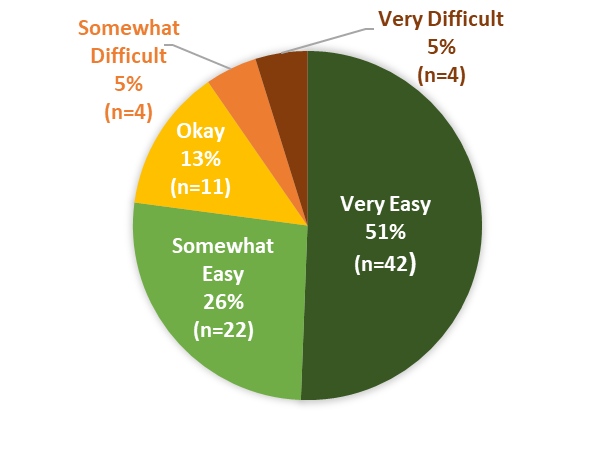

| **Participant Feedback** | **N=83** |
| --- | --- |
| Ease-of-Use |  |
| Very Easy | 42 (51%) |
| Somewhat Easy | 22 (26%) |
| Okay | 11 (13%) |
| Somewhat Difficult | 4 (5%) |
| Very Difficult | 4 (5%) |

### Table S1: Per-pathogen and overall concordance between detections for paired LRT samples (Biofire panel) and AveloMask samples (LightMix panel) stratified by study site.

Abbreviations: LRT, lower respiratory tract; OPA, overall percent agreement; PPA, positive percent agreement; NPA, negative percent agreement; PPV, positive predictive value; NPV, negative predictive value

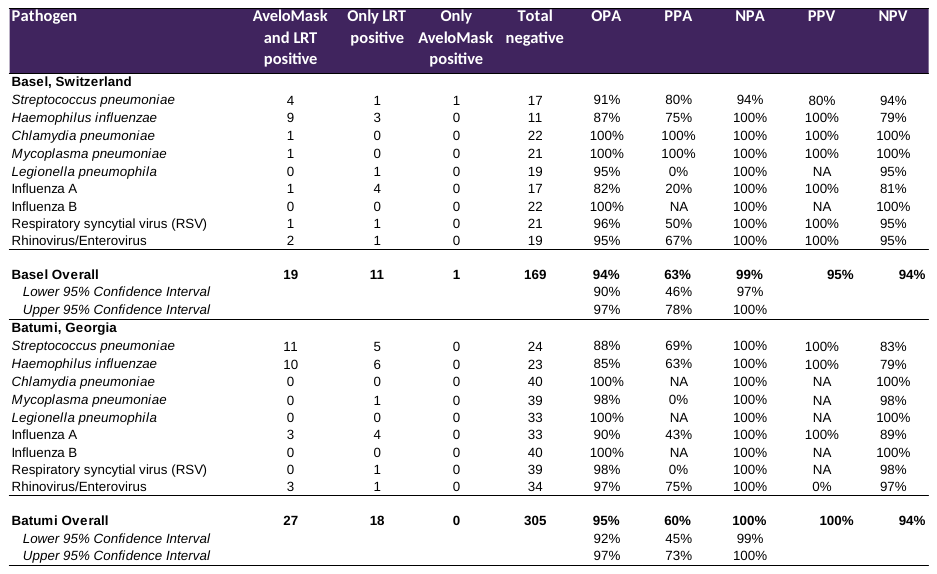

### Table S2: Per-pathogen and overall concordance between detections for paired LRT samples (Biofire panel) and AveloMask samples (LightMix panel) stratified by bacteria and virus.

Abbreviations: LRT, lower respiratory tract; OPA, overall percent agreement; PPA, positive percent agreement; NPA, negative percent agreement; PPV, positive predictive value; NPV, negative predictive value

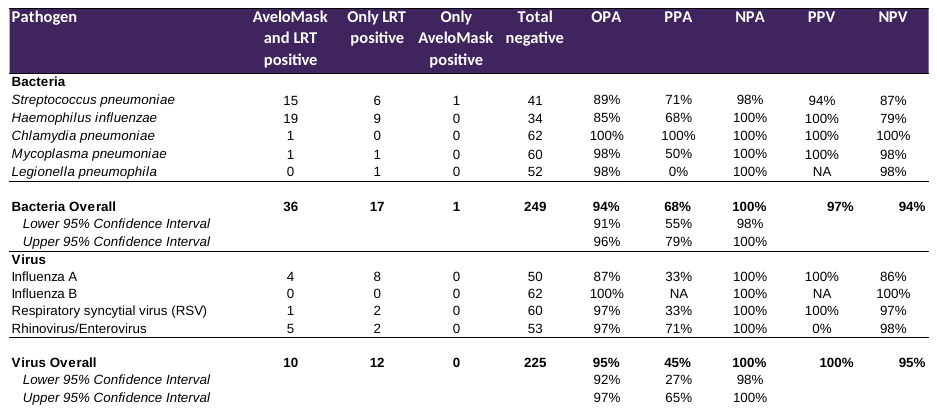

### Table S3: Per-pathogen and overall PPA for paired LRT samples (Biofire panel) and AveloMask samples (LightMix panel) stratified by LRT sample Ct value

Abbreviations: LRT, lower respiratory tract; PPA, positive percent agreement; Ct, cycle threshold

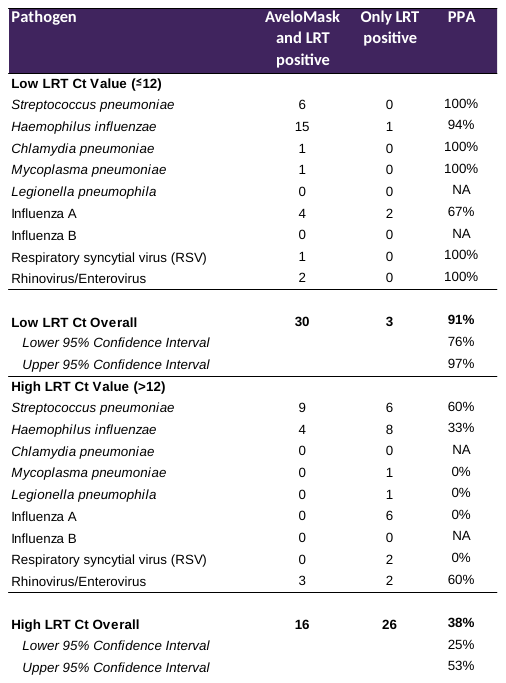

### Table S4: Per-pathogen and overall concordance between detections for paired LRT samples (Lightmix panel) and AveloMask samples (LightMix panel).

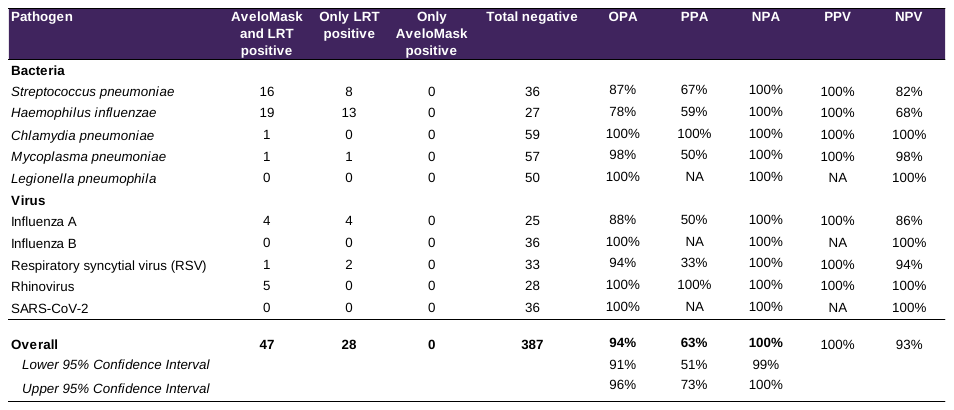
Abbreviations: LRT, lower respiratory tract; OPA, overall percent agreement; PPA, positive percent agreement; NPA, negative percent agreement; PPV, positive predictive value; NPV, negative predictive value

### Table S5: STARD Checklist

| **Section & Topic** | **No** | **Item** | **Done?** |
| --- | --- | --- | --- |
| **TITLE OR ABSTRACT** |  |  |  |
|  | **1** | Identification as a study of diagnostic accuracy using at least one measure of accuracy  (such as sensitivity, specificity, predictive values, or AUC) | yes |
| **ABSTRACT** |  |  |  |
|  | **2** | Structured summary of study design, methods, results, and conclusions  (for specific guidance, see STARD for Abstracts) | yes |
| **INTRODUCTION** |  |  |  |
|  | **3** | Scientific and clinical background, including the intended use and clinical role of the index test | yes |
|  | **4** | Study objectives and hypotheses | yes |
| **METHODS** |  |  |  |
| *Study design* | **5** | Whether data collection was planned before the index test and reference standard  were performed (prospective study) or after (retrospective study) | yes |
| *Participants* | **6** | Eligibility criteria | yes |
|  | **7** | On what basis potentially eligible participants were identified  (such as symptoms, results from previous tests, inclusion in registry) | yes |
|  | **8** | Where and when potentially eligible participants were identified (setting, location and dates) | yes |
|  | **9** | Whether participants formed a consecutive, random or convenience series | yes |
| *Test methods* | **10a** | Index test, in sufficient detail to allow replication | yes |
|  | **10b** | Reference standard, in sufficient detail to allow replication | yes |
|  | **11** | Rationale for choosing the reference standard (if alternatives exist) | yes |
|  | **12a** | Definition of and rationale for test positivity cut-offs or result categories  of the index test, distinguishing pre-specified from exploratory | yes |
|  | **12b** | Definition of and rationale for test positivity cut-offs or result categories  of the reference standard, distinguishing pre-specified from exploratory | yes |
|  | **13a** | Whether clinical information and reference standard results were available  to the performers/readers of the index test | yes |
|  | **13b** | Whether clinical information and index test results were available  to the assessors of the reference standard | yes |
| *Analysis* | **14** | Methods for estimating or comparing measures of diagnostic accuracy | yes |
|  | **15** | How indeterminate index test or reference standard results were handled | yes |
|  | **16** | How missing data on the index test and reference standard were handled | yes |
|  | **17** | Any analyses of variability in diagnostic accuracy, distinguishing pre-specified from exploratory | yes |
|  | **18** | Intended sample size and how it was determined | yes |
| **RESULTS** |  |  |  |
| *Participants* | **19** | Flow of participants, using a diagram | yes |
|  | **20** | Baseline demographic and clinical characteristics of participants | yes |
|  | **21a** | Distribution of severity of disease in those with the target condition | yes |
|  | **21b** | Distribution of alternative diagnoses in those without the target condition | no |
|  | **22** | Time interval and any clinical interventions between index test and reference standard | yes |
| *Test results* | **23** | Cross tabulation of the index test results (or their distribution)  by the results of the reference standard | yes |
|  | **24** | Estimates of diagnostic accuracy and their precision (such as 95% confidence intervals) | yes |
|  | **25** | Any adverse events from performing the index test or the reference standard | yes |
| **DISCUSSION** |  |  |  |
|  | **26** | Study limitations, including sources of potential bias, statistical uncertainty, and generalisability | yes |
|  | **27** | Implications for practice, including the intended use and clinical role of the index test | yes |
| **OTHER INFORMATION** |  |  |  |
|  | **28** | Registration number and name of registry | yes |
|  | **29** | Where the full study protocol can be accessed | no |
|  | **30** | Sources of funding and other support; role of funders | yes |
